# Tele-Rehabilitation for People with Visual Disabilities During COVID-19 Pandemic: Lesson Learned

**DOI:** 10.1101/2020.12.31.20249111

**Authors:** Suraj Singh Senjam, Souvik Manna, Praveen Vashist, Vivek Gupta

## Abstract

**Background:** The COVID-19 outbreak poses a global crisis in health care delivery system, including habilitation and rehabilitation services. In the present study, we shared our experiences on telerehabilitation services established primarily for students with visual disabilities (SwVD) amidst COVID-19 pandemic and its outputs.

**Methods:** Following the lockdown declared on March 23, 2020, the rehabilitative team of a tertiary eye center in north India received information that many VCS were stranded in schools for the blind in Delhi, and feeling with anxiety and panic in absence of teachers. Shortly, the room for vision rehabilitation clinic was set-up for tele-facilities. The intended services was explained while disseminating the mobile numbers. A semi-structured questionnaire consisting of closed and open-ended was developed to record COVID-19 knowledge and concerns. Inductive content analysis was used to report the qualitative information.

**Results:** As of June 30, 2020, a total of 492 clients contacted the team, with maximum from Delhi (41.5%), and predominantly males (78.8%). Around 80.3% of callers were VCS with age range of 11 to 30 years. The two most frequently encountered health needs were itching in eyes (36.1%) and headache (29%). Television news was the most used medium among callers to get COVID-19 information. Cough is a less frequently known mode of transmission (28%), similarly handwashing as a less known for prevention (17.2%). Eight concerns were recorded based on qualitative data analysis.

**Conclusion:** Tele-rehabilitation provides valuable insights and has the potential to address various concerns, uncertainty, anxiety, and fear among SVD during the pandemic.

## Introduction

The current Covid-19 pandemic has created unprecedented challenges not only in the provision of health care services but also habilitation and rehabilitation for persons with visual disabilities (PwVDs). The traditional way of practices like physical visits by patients or face to face consultation may pose with risk of exposure to the COVID-19 virus either from the providers to the patients or vice versa. As a result, there has been a global paradigm shift in delivering health care services using digital technologies in the form of telehealth or telemedicine or teleconsultation. ^[1, 2, 3]^ As of today, August 1, 2020, the natural history of the COVID-19, including its pathophysiology and scale of the route for transmission, though reported to be respiratory droplets, and long term health consequences have not been fully yet known. ^[4]^, On the other hand, the Government of India imposed many steps to limit the transmission of the virus, for example, complete or partial lockdown, closure of outpatients service of the hospitals, and transportations, etc. causing tremendous challenges in accessing healthcare services to public as well as disabled people.^[5, 6]^ In such unpredictable circumstances, the continued care for PwVDs in the form of telerehabilitation without increasing the risk of exposure to COVID-19 on healthcare or PwVDs is of overwhelming importance. Indeed, the continuation of services is critically important since PwVDs are likely to be more susceptible and may have serious health consequences if they contract with COVID-19.^[7]^

In general, vision rehabilitation service is not very urgent, but need a long interaction with patients, either in the form of counselling and education or one to one rehabilitation training with close contact. Since COVID-19 pandemic started in January 2020, an alternative way of providing rehabilitation service has been gaining a lot of attention in the form of telerehabilitation, besides teleconsultation using digital technologies across the world. ^[8]^ Telerehabilitation, unlike tele-medicine or tele-consultation in which medical consultation on diagnosis, treatment, and preventive practices are primarily involved, is an alternative strategy to help in accessing rehabilitation services using information and communication technologies amongst persons with disabilities (PwDs).^[9,10,11]^ Studies are reported on successful ophthalmic practices using tele-consultation facilities during the COVID-19 lockdown in India.^[12,13]^

Tele-rehabilitation for persons with disabilities, including visually challenged addresses, but not limited to, a range of habilitation and rehabilitation services that includes education, tele-counselling, consultation, monitoring, supervision, patient’s functional assessment, preventive measures, various rehabilitation therapy or intervention, training and demonstration to patients or caregivers as well as clinical management to a certain level, etc. ^[14,15]^ The communication between a disabled individual and rehabilitation professionals can be done with a variety of telecommunications platforms such a telephone, messaging, internet-based audio or videoconferencing, transferring of data to the rehabilitation professionals.^[8]^ The main advantages of telerehabilitation ensures theunnecessary travel to the hospital and maintain social distancing during the COVID 19 pandemic.

## Methodology

### How was the tele-rehabilitation service conceived amidst the COVID-19 pandemic?

Shortly following the nationwide emergency lockdown, declared on March 23, 2020, the team of Vision Rehabilitation &Training Center (VR&TC) of a tertiary eye care centre in north India noticed that many SwVD were stranded at schools for the blind (residential) or vocational training centers in National Capital Regions, India. The team immediately contacted and discussed with the authorities of few leading schools for the blind and Non-Government Organizations (NGOs) about the need of telerehabilitation or teleconsultation during the pandemic lockdown. We also discussed the feasibility of telerehabilitation services, accessibility of smartphones among students or so, and the prospects of services. The team did a trial with a few students and took the feedback about the need for such an initiative during the pandemic. On April 20, 2020, tele-rehabilitation was started and shared the contact numbers to the representative of NGOs and schools. We shared the experiences that we learned from the tele-rehabilitation services that was set up for SwVD during the COVID-19 lockdown in the present paper.

### Setting up and equipment for tele-rehabilitation

The team used three smartphones and two tablets for the services. Audio based, VoIP (Voice over Internet Protocol like WhatsApp calling, online Zoom platform), and messaging were the various communication platforms that were used for communication. A dedicated room used for vision rehabilitation before the pandemic was designed for the purpose. Mobile numbers, the details of working hours for consultation were disseminated to the authorities of various organizations. A rehabilitation team primarily consisting of faculty members, a Ph.D. scholar with qualification in MBBS, MD, two rehabilitation staff who have been working for the last 5 years were involved in the telerehabilitation. The team aimed to receive all calls from clients. The term ‘clients’ indicated all recipients of telerehabilitation services irrespective of whether callers are beneficiaries (disabled individuals) or caregivers or parents or family members of disabled child. All callers may not be beneficiaries. For example, if a parent (caller) called the team for his or her children having retinopathy of prematurity (beneficiary). If the team missed any call, a call back was done to the same number. Follow up calls were also made to know the status or whereabout of PwVDs.

The faculty who looks after vision rehabilitation educated and trained for a day to all the team about COVID-19 information, e.g. cause, symptoms, mode of transmission, etc., and preventive measures along with techniques using various sources like The Centre for Diseases Control and Prevention, The Department of Health and Human Services, USA; The World Health Organization and Ministry of Health and Family Welfare, Govt. of India, etc. Using these sources will help to provide the scientific and reliable information about COVID-19. The training also included about rapport building of the clients and filling out the form. These educational materials were made available in the tele-room for ready references.

### Intended services in tele-rehabilitation

In the present telerehabilitation services, we did not intend to provide all the components since it was not feasible during the emergency COVID-19 lockdown, rather aimed for education and counselling, monitoring if any VCS visits to the centre earlier, problem-solving communication to clients’ uncertainty, training and demonstration about preventive measures to caregivers or family members as well as supportive supervision during the lockdown period. We also aimed to facilitate healthcare access through the hospital telemedicine and emergency services if required as well as management of some minor ailments as per the Govt. of India’s telemedicine guidelines. If any caller seeking for further health care, including ocular problems, not only we advised to contact the concerned hospital teleconsultation services, but also facilitate to avail the tele-facilities or emergency healthcare.

### Study tool

A semi-structured questionnaire was used for the study that consisted of closed and open-ended questions (Annexure 1). An example of open-ended was “what are your concerns or worries during COVID-19 pandemic? The information received was written on the space provided in the tool. A remark or action taken if required was also present. Each caller was explained about the information that we would like to obtain during the tele-rehabilitation. Since, it was not possible to have a written consent, an implied consent of the callers was considered as the call was initiated from the clients. We relied on the information provided by the clients.

The tool was pretested to five initial clients to standardize and made necessary changes thereafter. While developing the tool, the following steps were followed to standardize it. Firstly, we reviewed for any existing similar tool, 2^nd^-develop an initial draft in consultation with other experts (faculty members), 3^rd^-the same was pre-tested to non-study clients; 4^th^-checking the languages, wording, clarity, easy or difficult to understand, including layout; 5^th^-input all coding and finalized it. For instance, during the process we removed the question on cause of visual impairment, or avoided the address detailed or revised the list of common ailments.

### Data analysis

Since we used semi-structured recording forms for the study, the quantitative information was presented descriptively, e.g. frequency, percentage, calculated using STATA-15 (StataCorp.2015.Stata Statistical Software: Release 15. College Station, Tx: StataCorp LP), whereas inductive content analysis was done for the qualitative information based on Thomas’s method manually. ^[16]^ Inductive analysis was performed to gain meaningful information received or to understand more insights. The process consists of five steps. Firstly, initial reading of text data or information; secondly, identification of specific text segments relevant to objectives; third, labeling the segments of the text to create categories or theme; fourth, reducing overlap and redundancy among categories, and fifth, creating a model incorporating most important categories. A total of three researchers have participated in the process of analysis. For trustworthiness and credibility, a series of debates among researchers as well as consistency checking was done in case of disagreement in the analysis process. To further ensure the trustworthiness, “the quotes” from the callers/beneficiaries are reported in the results.

## Results

As of June 30, 2020, a total of 492 calls were received from different parts of the country with a maximum from Delhi (41.5%) followed by Uttar Pradesh (23.6%). The male callers were 78.8%, nearly two-thirds of them owned a smartphone, and 43.3% of callers/beneficiaries made the call without any assistant. Around 96% of the callers belonged to visually disabled, with 16.5% of the callers’ visual status were unknown (waiting or applied for certificates). The majority of callers/beneficiaries belonged to age group between 11 to 30 years (82.3%) and were students (80.3%, Table 1).

**Table 1:** Characteristics of callers/beneficiaries (N=492)

| **Characteristics** | **No. of callers/beneficiaries (n)** | **Percent (%)** |
| --- | --- | --- |
| Location of the callers |  |  |
| Delhi | 204 | 41.5 |
| Uttar Pradesh | 116 | 23.6 |
| Bihar | 59 | 11.9 |
| Haryana | 55 | 11.2 |
| Jharkhand | 18 | 3.7 |
| Rajasthan | 14 | 2.8 |
| Madya Pradesh | 11 | 2.2 |
| Other states | 15 | 3.1 |
| Gender of the callers |  |  |
| Male | 388 | 78.8 |
| Female | 104 | 21.2 |
| Own a smartphone by disabled/beneficiaries |  |  |
| Yes | 280 | 56.9 |
| No | 212 | 43.1 |
| Assistance in dialing |  |  |
| Beneficiaries (Disabled) self | 213 | 43.3 |
| Assisted dial/caregivers/team | 279 | 56.7 |
| Degree of disability of the callers/beneficiaries |  |  |
| Waiting for certificate | 81 | 16.5 |
| <40% | 36 | 7.3 |
| ≥40% | 375 | 76.2 |
| Type of disabilities of the callers/beneficiaries |  |  |
| Visual | 472 | 95.9 |
| Physical | 14 | 2.9 |
| Mental | 5 | 1.0 |
| Deaf-mute | 1 | 0.2 |
| Caregivers |  |  |
| Self & independent | 227 | 46.1 |
| Family members/parents | 218 | 44.2 |
| Peers/other caretakers (hostel) | 47 | 9.7 |
| Age of the beneficiaries (disabled) |  |  |
| 0-10 years | 33 | 6.7 |
| 11-17 years | 208 | 42.3 |
| 18-30 years | 197 | 40.0 |
| 31-59 years | 51 | 10.4 |
| 60 years and above | 3 | 0.6 |
| Occupation of the beneficiaries (disabled) |  |  |
| Students | 395 | 80.3 |
| Pre-school | 19 | 3.8 |
| Govt. job | 5 | 1.0 |
| Shopkeepers/Pvt works/Special NGOs | 23 | 4.8 |
| Homemakers | 9 | 1.8 |
| Others/Unemployed | 41 | 8.3 |

### Presenting complaints or seeking for health care from callers or beneficiaries

Besides the various concerns on COVID-19 pandemic, around 47.9% of clients (235) reported physical health problems with a total complained of 335 among beneficiaries The three most frequently presenting complaints were itching in the eyes (36.1% of overall complaints), headache (29.0% of complaints) and watering in eyes (16.1%, Table 2). The minimum complaints received from callers was redness in the eyes (1.5% of the total).

**Table 2:**
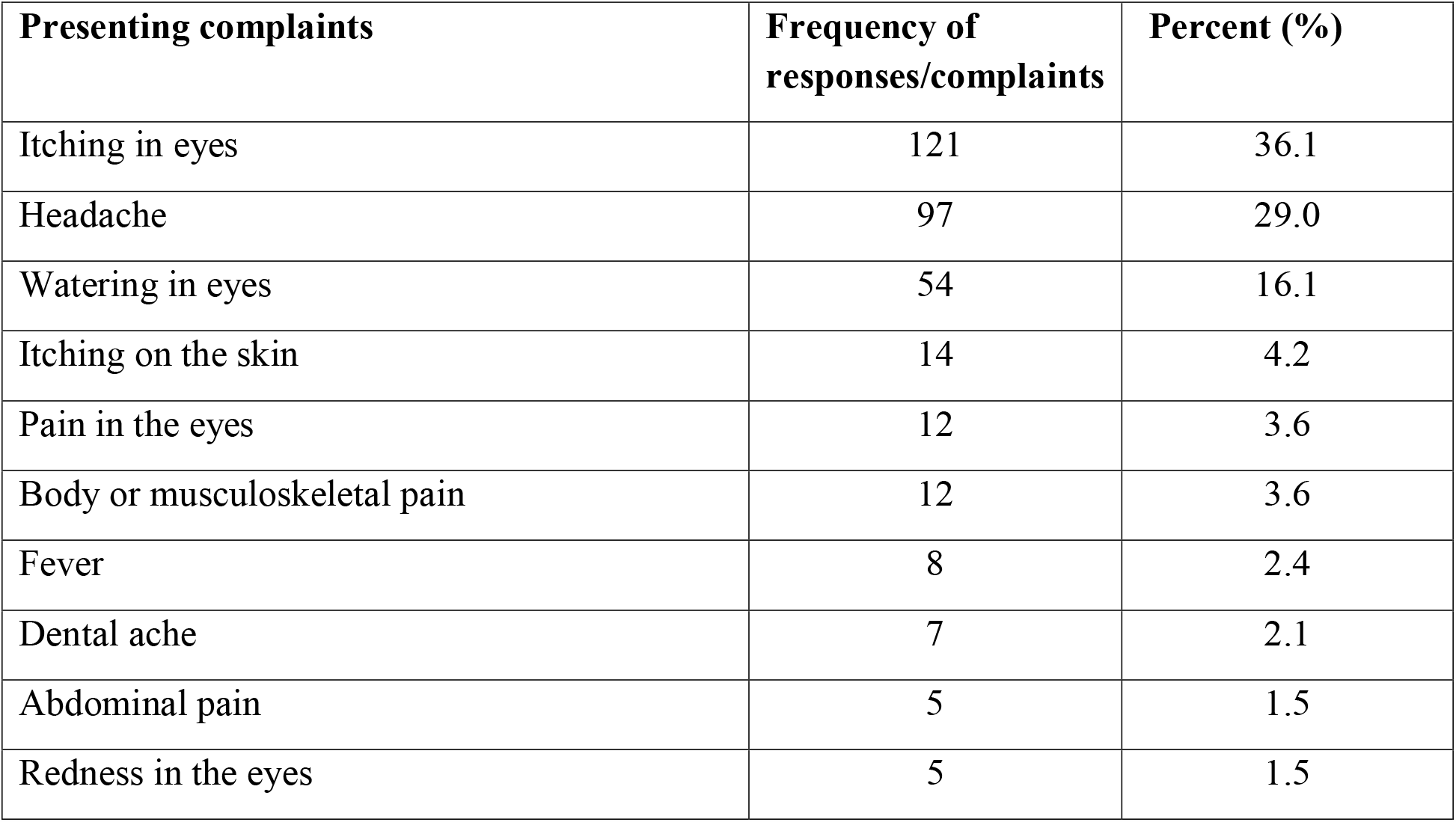
Frequency of presenting complaints or seeking healthcare (responses) made by the callers or beneficiaries.

| **Presenting complaints** | **Frequency of responses/complaints** | **Percent (%)** |
| --- | --- | --- |
| Itching in eyes | 121 | 36.1 |
| Headache | 97 | 29.0 |
| Watering in eyes | 54 | 16.1 |
| Itching on the skin | 14 | 4.2 |
| Pain in the eyes | 12 | 3.6 |
| Body or musculoskeletal pain | 12 | 3.6 |
| Fever | 8 | 2.4 |
| Dental ache | 7 | 2.1 |
| Abdominal pain | 5 | 1.5 |
| Redness in the eyes | 5 | 1.5 |

### Awareness and knowledge about COVID-19 pandemic

A total of 722 responses from 491 callers were recorded when we asked what media help to get the information on COVID-19 disease. Television news is the most frequently reported medium to receive about the pandemic (59.4% of all responses) and followed by friends or peers (9.6%) and using mobile internet data (7.9%, Table 3). When we enquired about the mode of transmission of disease, transmission by the contact or touch is the most frequent response (36.4%; 408 out of 1122), followed by sneezing (35.2% of total responses, Table 3). In the context of preventive strategies from COVID 19 diseases, social distancing (41.6%; 470 out of 1136) and wearing a face mask (41.2%) were frequent responses from the callers.

**Table 3:**
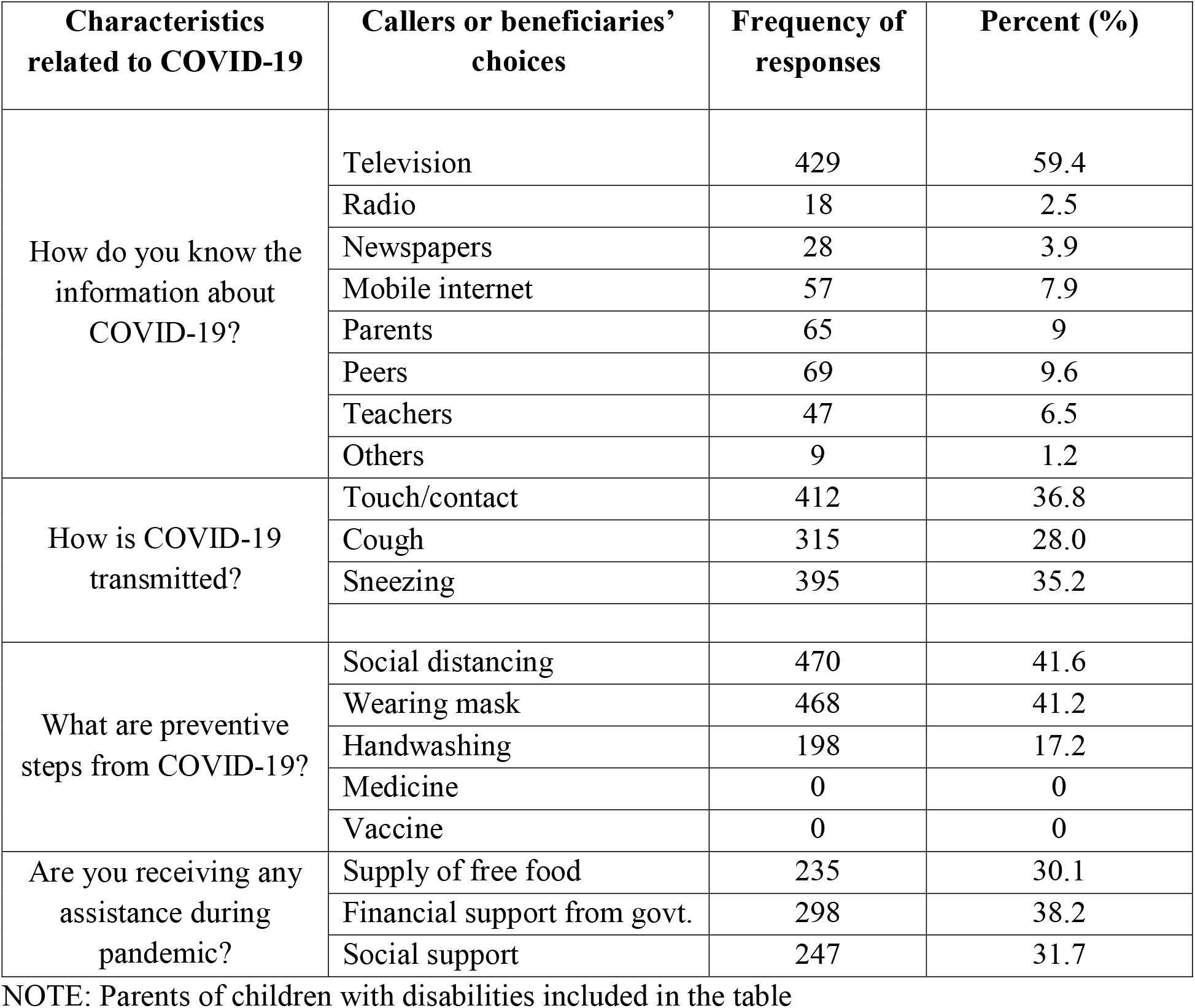
**Perceived awareness and knowledge about COVID-19 among callers or beneficiaries**

### Findings from the inductive content analysis

The intent of inductive content analysis (ICA) was to gain insights of qualitative information based on the open-ended question on concerns or worries due to the pandemic. We recorded responses from 290 consecutive callers about their concerns and problems faced during the COVID −19 pandemic. This optimum sample size was guided by the degree to which incoming qualitative information adequately answer the questions.^[17]^ So, we avoided further recording of the concerned information because there was no or a little newly added meaningful information (data saturation). Such data saturation for estimating sample size is a valid method in collecting qualitative information.^[18]^ The following various categories were revealed and were facilitated through the Tele-rehabilitation to clients. There were eight themes or categories that emerged: 1) The COVID-19 disease related, 2) The lockdown related, 3) Health-related, 4) Livelihood related, 5) Education related, 6) Social related, 7) Certificates related, and 8) Empowerment related (Table 4).

**Table 4:** **Callers or beneficiaries’ (visually disabled) concerns based on inductive content analysis**

| **Themes /categories** | **Description of concerns** | **Quotes** |
| --- | --- | --- |
| COVID-19 pandemic related | Lack of Information about the COVID-19 Pandemic. | *“I heard the disease, but not sure what is all about COVID 19 disease?”* |
|  | Feeling anxiety, fear about the COVID-19 disease. | *“After listening COVID 19 from the news, I am feeling nervous and fear, we are not even meeting each other in the schools”.* |
|  | Worries, apprehension, about safety and prevention from the COVID-19 disease. | *“Not sure how to maintain social distancing and how long can we wear face masks”* |
|  | Worries about food safety, buying foods groceries from the markets |  |
| Lockdown related | Unable to go their native town or village because of closure transportations. | *“We (students in schools for the blind) want to go home and stay with parents, but due to lockdown we are not able to commute”* |
|  | Could not find accommodation during the lockdown. | *I’ve come to Delhi for vocational training course, but neither I could get admission nor place to stay now”* |
|  | Reluctance to stay at local/villages government quarantine facilities. | *“I’ve reached my home safely but was being told to stay at village quarantine centre, I’m scare of staying there.”* |
| Health issues | Worries for an emergency health care access. | *“I’m staying in rented house, if I’ve some medical problem, I’m not sure where to go.”* |
|  | Uncertainty in follow up for their eye problems. | *“I’m not able to visit hospital for follow up examination”* |
|  | Worries about health seeking for common health problems. |  |
| Empowerment issues | Inaccessible about government smartphone applications, disabled benefit schemes, e.g. Aarogya Setu App, food supply.<br>Not having any accessible contact numbers for disabled or government accessible platform for help | *“I’ve heard about the Aarogya Setu App launched by Government, but it is not accessible”.*<br>*“We need online platform which is accessible for daily activities “* |
| Education related | Concerns about the admission to schools for blind schools, college admission.<br>Worries on crossing the age limits for the admission. | *“I need to enroll my son to the special school for his education, I’m worried about his age limits”*<br>*“I could not write one or two board papers due* |
|  | Not able to visit schools for the blind for admission.<br>Worries and anxiety on the board papers left to write and impact on future admission. | *to sudden nationwide lockdown, now I'm very much worried what will be future outcome"*<br>*"I don't know what will happen in my next admission"* |
| Livelihood issues | Apprehension on completion of vocational training, and livelihood activities training.<br>Worries about applying loan, and future Job.<br>Cannot apply for job due to old certificates.<br>Fear of losing their job. | *"I still have to complete my vocational training course, but now I could not complete, I cannot apply for the job or government loan"*<br>*"The organization asked me to submit a new disability certificate, they don't accept old certificate, so I cannot apply for renewal"* |
| Social issues | People reluctant to help in moving out, e.g. crossing the road.<br>Facing social stigma & discrimination while outdoor movement during the pandemic. | *"I hardly find anyone who can help in crossing the road"*<br>*"I go for shopping to buy ration and fruits, but shopkeeper doesn't treat me well, saying I might have touch everywhere to come here, so asked me stay away far of".* |
| Certificate related | Waiting for issuance of disability certificate.<br>Require a new certificate for government scheme applications.<br>Not able to renew of visual disability certificates<br>Unable to apply for certificates | *"I've to apply for disability certificate for my daughter school admission, I'm very worried when should I am able to apply"* |

### Services and assistance

Clients were educated about COVID-19: sign and symptoms, mode of transmission, potential source of sites or surfaces, preventive measures like wearing of face masks, handwashing. Counselling was actively done to reduce the anxiety and panic felt; follow up calls were made as a part of monitoring; online audio-video based teaching or demonstration on technique of handwashing, wearing of masks; management of some minor ailments as per the Govt. of India guidelines; group meeting using online platform, e.g. Zoom; and yoga sessions were a part of the tele-rehabilitation services. The team assisted in accessing healthcare needs either through the hospital tele-medicine or informed the clients about the emergency facilities available with a subsequent communication to patients who had been facilitated earlier. For example, the team recontacted a SwVD with a severe pain abdomen who was facilitated to access an emergency services during the lockdown.

There were services for problem-solving that the team had never thought of, for instance, few visually challenged students/persons who came to Delhi for a new disability certificates or for renewal or vocational training were not able to get a place to stay during the lockdown, our team coordinated with available residential vocational training centers or schools, and arranged a place to stay for temporarily; advised to family members and village leaders to allow for home isolation rather than village quarantine facilities for SwVDs who returned to their respective villages; even the team helped SwVDs to locate misplaced households items with smartphone applications, etc.

## Discussion

The world has been facing a crisis on health care delivery system since the WHO announced the pandemic of COVID-19 disease in March 11, 2020, with over 35.2 million confirmed cases and over a million deaths from 235 countries as of October 5, 2020.^[19]^ Following this announcement, the routine conventional face to face healthcare practices suffers a great disruption, and suddenly gaining due attention with telehealth practices employing user friendly electronic communication technologies. Such tele-health services was shown to be useful in previous outbreaks like SARS-COV and MERS-COV, Ebola and Zika viruses. ^[20]^

Among various modalities of telehealth, tele-rehabilitation has been shown to be useful in morbidities like stroke, diabetes, cancer, neurological problems, patients with cardiac or musculoskeletal problems. ^[21,22,23,24]^Tele-rehabilitation for visual impairment (low vision and blind) can be considered as a subset of under the umbrella tele-rehabilitation services. The goal of vision rehabilitation whether in person or remotely via tele-rehabilitation, is to improve visual functioning, and daily living activities, including instrumental activities as well as social and psychological well-being among PwVDs. Evidence exists that the usefulness of such tele-(vision) rehabilitation services in improving the overall life functioning and quality of life among people with low vision and blindness.^[10]^ However, the present telerehabilitation established during the COVID-19 emergency lockdown is primarily to alleviate the fear and panic felt among SwVDs. Though the team could not provide all components of tele-rehabilitation, a wide range of feasible services, including training and demonstration of COVID-19 preventive measures are covered. The team also helps in solving previously unexpected problems for PwVDs during the lockdown. Traditionally, the rehabilitation services is delivered to PwDs by a multidisciplinary team of trained personnel. Similarly, the present tele-(vision) rehabilitation service is involved a team of trained staff of a tertiary eye care hospital with SwVDs as target audiences.

The results show that most of the callers are students (80.3% of the total) with male predominant (78.8%). This finding corresponds to what teleconsultation services intended. The male predominance among callers could be because in most of these schools for the blind are predominantly occupied by male students than their female counterparts. Other factors could be female students with visual disabilities are not having smartphones or reluctance to call a male rehabilitation staff, since we do not have a female rehabilitation staff. Around 56.9% of visually disabled have a smartphone and 43.3% of beneficiaries dialed without any assistant (Table 1). This information is important because these group of disabled are the potential candidates for training on use special smartphone applications for visual impairment (VI) through tele-rehabilitation in the futureand study on the utilities of such applications. This gives an opportunity for further services and research.

The result also indicates that around 16.5% of the callers do not have visual disability certificates or not able to renew it or applied but not received yet. This finding is a concern because such visually disabled cannot avail of various benefits without disability certificates provided by the government during lockdown periods. For example, the Government of Delhi ensures financial security to PwDs having disability certificates to the double amount during the lockdown.^[25]^

The team also received calls for health-related issues as a primary reason. The two most frequently presenting eye or health problems were itching in the eyes and headache. These findings are different from other teleconsultation studies reported from the north and south India in which redness, blurring of vision, pain, and watering were common complaints. ^[13]^ The difference could be on account of difference in morbidities since the callers or beneficiaries in the present study are the population with a visual disability. The next frequent complaint was headache among callers. Perhaps the headache may be related to the extensive use of smartphones. An individual with a visual loss is more likely to use a mobile phone for a longer time than people without a visual loss. Very few studies reported that a nonspecific of ill-health like a headache is due to the effects of the radiofrequency electromagnetic field from a mobile phone.^[26,27]^

Since the callers in present study already have a visual disability, they are likely to use the smartphone for audio purposes, not for screen exposure to the eyes. Therefore, a further study is warranted on whether the headache accounts due to exposure to the electromagnetic field emitted by smartphone, not by screen exposure to the eyes.

The news on the Television channels is the most used medium among callers to get COVID-19 information. Since television is ubiquitous in every family or organization and the most common medium to get information among PwVDs, it can also be a medium to reduce the stigma, anxiety, and panic conceived during the pandemic. However, an accessible and appropriate information system should widely be available that helps in reducing the fear felt among PwVDs during the outbreak. Very few responses are received handwashing as a preventive measure among the callers. They need to be educated about handwashing with appropriate techniques as it is one of the best preventive steps from the COVID-19 virus.

Table 4 shows that not only concerns of PwVDs about COVID-19, but also they worry about other aspects of lives during the pandemic, for example, feeling of anxiety and fear while buying foods or groceries items, challenges in finding accommodation during the lockdown, the compounded social stigma against them or the fear of losing job, etc.

During such a pandemic followed by a severe health care delivery crisis, visually disabled are needed to be informed and educated about COVID -19 along with various preventive strategies like need of changing behavior, social distancing, avoid public gathering, wearing of face or medical masks, avoiding unnecessary touch, etc. A platform in the form of tele-(vision) rehabilitation in every eye care facility across the country is required so that PwVDs can obtain the necessary awareness and knowledge about the COVID-19.

Performing a regular yoga sessions through online platforms, connecting with friends and family members, and rehabilitation professionals, sharing experiences and help to each other among visually disabled, conducting online group meeting, avoid listening news channels obsessively are some of the useful activities to reduce anxiety fear and surrounding uncertainty during the COVID-19 pandemic. The team organized such activities as a part of remedy. Obtaining the information only from trustworthy and reliable sources such the government website, rehabilitation professionals, and the World Health Organization is critically important to avoid any misinformation. Not only this, PWVDs should be learned about food safety, handling daily groceries items though tele-(vision) rehabilitation services.

## Limitations

The study has few limitations. Neither we could do any physical verification of associated disability nor assessed the degree of disability. We relied on the caller’s response through phone for their type and degree of disability. Around 16.5% of callers who are not having certificates that the team could not get information on disabilities. As a service limitation, we could not provide all components of rehabilitation which are potentially feasible through tele-facilities.

## Conclusions

Since the COVID-19 pandemic declared by the WHO on March 11, 2020, a global paradigm shift in health care practices using information and communication technologies has been gaining a lot of attention. While the conventional way of communicative health care practices, i.e. direct face to face contact, poses a risk of exposure between providers and patients, tele-(vision) rehabilitation offers a safe and an efficient way of providing all reliable information of COVID-19, including various preventive strategies among visually disabled. The platform also helps in psychological counselling for fear and panic, facilitating and addressing the many unseen challenged faced by visually disabled. However, many developing countries, including India, lack a regulatory framework to establish such digital services into the health care system. Based on **t**he experiences we presented here, tele-(vision) rehabilitation particularly for people visual disabilities can be considered as an essential service wherever vision rehabilitation clinic exists.

## Data Availability

All final data is enclosed in the manuscript.

## References

1. Golinelli D, Boetto E, Carullo G, Nuzzolese AG, Landini MP, Fantini MP. How the COVID-19 pandemic is favoring the adoption of digital technologies in healthcare: a literature review. [Last accessed on July 21,2020];Available from: https://www.medrxiv.org/content/10.1101/2020.04.26.20080341v2.full.pdf

2. Nair AG, Gandhi RA, Natarajan S. Effect of COVID-19 related lockdown on ophthalmic practice and patient care in India: Results of a survey. Indian J Ophthalmol 2020;68(5):725–30.

3. Ohannessian R, Duong TA, Odone A. Global Telemedicine Implementation and Integration Within Health Systems to Fight the COVID-19 Pandemic: A Call to Action. JMIR Public Heal Surveill 2020 [Laset accessed on July 28, 2020];6(2):e18810. Available from: /pmc/articles/PMC7124951/?report=abstract

4. Rismanbaf A. Potential Treatments for COVID-19; a Narrative Literature Review. Arch Acad Emerg Med 2020;8(1):e29. Available from: http://www.ncbi.nlm.nih.gov/pubmed/32232214

5. Khanna RC, Cicinelli MV, Gilbert SS, Honavar SG, Murthy GVS. COVID-19 pandemic: Lessons learned and future directions. Indian J. Ophthalmol.2020;68(5):703–10.

6. Ministry of Health & Family Welfare Governtment of India. COVID-19 India. [Last accessed on May 15, 2020];Available from: https://www.mohfw.gov.in/

7. Senjam SS. Impact of COVID-19 pandemic on people living with visual disability. Indian J Ophthalmol 2020;68(7):1367–70.

8. Leochico CFD. Adoption of telerehabilitation in a developing country before and during the COVID-19 pandemic. Ann Phys Rehabil Med 2020, June 13; doi: 10.1016/j.rehab.2020.06.001 [Epub ahead of print]

9. Christy B, Keeffe J. Telerehabilitation during COVID-19: Experiences in service delivery from South India. Indian J. Ophthalmol.2020;68(7):1489–90.

10. Bittner AK, Yoshinaga PD, Wykstra SL, Li T. Telerehabilitation for people with low vision. Cochrane database Syst Rev 2020;2(2):CD011019. Available from: https://pubmed.ncbi.nlm.nih.gov/32102114/

11. Brennan DM, Tindall L, Theodoros D, Brown J, Campbell M, Christiana D, et al. A blueprint for telerehabilitation guidelines--October 2010. Telemed J E Health 2011;17(8):662–5. Available from: https://pubmed.ncbi.nlm.nih.gov/21790271/

12. Das A V., Rani PK, Vaddavalli PK. Tele-consultations and electronic medical records driven remote patient care: Responding to the COVID-19 lockdown in India. Indian J Ophthalmol 2020;68(6):1007–12.

13. Pandey N, Srivastava RM, Kumar G, Katiyar V, Agrawal S. Teleconsultation at a tertiary care government medical university during COVID-19 Lockdown in India - A pilot study. Indian J Ophthalmol 2020;68(7):1381–4.

14. Brennan D, Tindall L, Theodoros D, Brown J, Campbell M, Christiana D, et al. A Blueprint for Telerehabilitation Guidelines. Int J Telerehabilitation 2010;2(2):31–4.

15. Zampolini M, Todeschini E, Guitart MB, Hermens H, Ilsbroukx S, Macellari V, et al. Tele-rehabilitation: Present and future. Ann. Ist. Super. Sanita 2008;44(2):125–34.

16. Thomas DR. A General Inductive Approach for Analyzing Qualitative Evaluation Data. Am J Eval 2006;27(2):237–46.

17. Prscilia R, Robinson ET, Tolley EE. Qualitative Methods in Public Health: A Field Guide for Applied Research 2005, JOSSEY-BASS, San Francisco, ISBN 0-7879-7634.

18. Guest G, Namey E, Chen M. A simple method to assess and report thematic saturation in qualitative research. PLoS One 2020;15(5):e0232076. Available from: https://dx.plos.org/10.1371/journal.pone.0232076

19. The World Health Organization. Coronavirus disease 2019. [Last accessed on Oct. 5, 2020 May 2];Available from: https://www.who.int/emergencies/diseases/novel-coronavirus-2019

20. Ohannessian R, Duong TA, Odone A. Global Telemedicine Implementation and Integration Within Health Systems to Fight the COVID-19 Pandemic: A Call to Action. JMIR Public Heal Surveill 2020;6(2):e18810.

21. Peretti A, Amenta F, Tayebati SK, Nittari G, Mahdi SS. Telerehabilitation: Review of the State-of-the-Art and Areas of Application. JMIR Rehabil Assist Technol 2017;4(2):e7. Available from: /pmc/articles/PMC5544892/?report=abstract

22. Laver KE, Adey-Wakeling Z, Crotty M, Lannin NA, George S, Sherrington C. Telerehabilitation services for stroke. Cochrane Database of Systematic Reviews 2020, Issue 1. Art. No.: CD010255. DOI: 10.1002/14651858.CD010255.pub3

23. Sahu D, Rathod V. Letter to the Editor regarding Menendez et al: “Orthopedic surgery post COVID-19: an opportunity for innovation and transformation”. J Shoulder Elbow Surg. 2020 Oct;29(10):1957–1958. doi: 10.1016/j.jse.2020.07.001. Epub 2020 Jul 10. PMID: 32659460; PMCID: PMC7351064.

24. Scherrenberg M, Wilhelm M, Hansen D, Völler H, Cornelissen V, Frederix I, et al. The future is now: a call for action for cardiac telerehabilitation in the COVID-19 pandemic from the secondary prevention and rehabilitation section of the European Association of Preventive Cardiology. Eur J Prev Cardiol. 2020 Jul 2:2047487320939671. doi: 10.1177/2047487320939671. Epub ahead of print. PMID: 32615796.

25. Delhi government raises widows, senior citizens, disabled pensions - The Financial Express. [Last accessed on Aug. 3, 2020];Available from: https://www.financialexpress.com/economy/delhi-government-raises-widows-senior-citizens-disabled-pensions/551365/

26. Wang J, Su H, Xie W, Yu S. Mobile Phone Use and The Risk of Headache: A Systematic Review and Meta-analysis of Cross-sectional Studies. Sci Rep 7, 12595 (2017). https://doi.org/10.1038/s41598-017-12802-9

27. Röösli M, Frei P, Mohler E, Hug K. Systematic review on the health effects of exposure to radiofrequency electromagnetic fields from mobile phone base stations. Bull World Health Organ 2010;88(12):887–96.

